# The challenge of controlling an auditory BCI in the case of severe motor disability

**DOI:** 10.1101/2023.01.10.23284295

**Authors:** Perrine Séguin, Emmanuel Maby, Mélodie Fouillen, Anatole Otman, Jacques Luauté, Pascal Giraux, Dominique Morlet, Jérémie Mattout

## Abstract

**Background:** The locked-in syndrome (LIS), due to a lesion in the pons, impedes communication. This situation can also be met after some severe brain injury or in advanced Amyotrophic Lateral Sclerosis (ALS). In the most severe condition, the persons cannot communicate at all because of a complete oculomotor paralysis (Complete LIS or CLIS). This even prevents the detection of consciousness. Some studies suggest that auditory brain-computer interface (BCI) could restore a communication through a « yes-no » code.

**Methods:** We developed an auditory EEG-based interface which makes use of voluntary modulations of attention, to restore a yes-no communication code in non-responding persons. This binary BCI uses repeated speech sounds (alternating “yes” on the right ear and “no” on the left ear) corresponding to either frequent (short) or rare (long) stimuli. Users are instructed to pay attention to the relevant stimuli only. We tested this BCI with 18 healthy subjects, and 7 people with severe motor disability (3 “classical” persons with locked-in syndrome and 4 persons with ALS).

**Results:** We report online BCI performance and offline event-related potential analysis. On average in healthy subjects, online BCI accuracy reached 86% based on 50 questions. Only one out of 18 subjects could not perform above chance level. Ten subjects had an accuracy above 90%. However, most patients could not produce online performance above chance level, except for two people with ALS who obtained 100% accuracy. We report individual event-related potentials and their modulation by attention. In addition to the classical P3b, we observed a signature of sustained attention on responses to frequent sounds, but in healthy subjects and patients with good BCI control only.

**Conclusions:** Auditory BCI can be very well controlled by healthy subjects, but it is not a guarantee that it can be readily used by the target population of persons in LIS or CLIS. A conclusion that is supported by a few previous findings in BCI and should now trigger research to assess the reasons of such a gap in order to propose new and efficient solutions.

**Clinical trial registrations:** N° NCT02567201 (2015) and NCT03233282 (2013)

## Introduction

The typical locked-in syndrome (LIS) is caused by a lesion in the pons, and patients are able to communicate only with movements of their eyes or eyelids [1]. This condition of total paralysis can also be encountered in the amyotrophic lateral sclerosis (ALS). In a completely locked-in state (CLIS), the person cannot communicate at all, which implies that the diagnosis of the state of consciousness is clinically impossible or very delayed [2]. In general, the assessment of consciousness in non-responsive patients remains challenging, with up to 40% of patients in minimal conscious state that may be misdiagnosed in a vegetative state by non-expert teams [3]. Even after a careful behavioral assessment, there is still the possibility that the patient cannot show any response to command because of complete motor impairment. The development of paraclinical assessments of patients with disorders of consciousness revealed that some of them, although diagnosed in a vegetative state or even in coma [4], [5], were able to prove their consciousness by willfully modulating their brain activity (command following) when they were asked to, and thus should be considered as in a complete locked-in state. The first striking demonstration of such a cognitive motor dissociation (dissociation between awareness and motor capacity) was reported in 2006, using fMRI [6].

EEG-based Brain-Computer Interfaces (BCI) are promising tools to detect a cognitive motor dissociation [7]. Indeed, they measure brain activity directly, in real-time, and enable repeated assessments at the patient’s bedside. Furthermore, they may also be used as communication devices. However, restoring communication with these patients once the diagnosis of command-following has been made, remains a major issue. The authors of two studies published in 2017 claimed that a communication was restored with people in CLIS [8], [9], but some flaws have been observed in their methodologies and their results remain controversial [10], which led to the retraction of one of them [11]. In another study [12], the authors used a steady state visually evoked potential BCI, which they evaluated longitudinally over 27 months in a patient with ALS. This patient could train with the BCI during three months before entering a CLIS state. The reliability of the BCI proved to be fluctuant, with accuracies below chance level in 13 out of 40 sessions [12]. A recent publication with an implanted intra-cortical electrode in the dominant left motor cortex demonstrated both the feasibility and the striking limitations of communication with a CLIS patients at the advanced stage of ALS [13]. This patient was implanted once he was already in a CLIS state with no residual eye movements, as attested by EOG. During the first stages of training, it appeared that when the patient was instructed to attempt or imagine hand, tongue or foot movements, no cortical response could be detected. Reliable yes-no responses were finally obtained three months after implantation thanks to a neurofeedback protocol. Tones with two different frequencies were provided according to the neural activity. During the 356 days following this training paradigm, he obtained an accuracy of 86.6 % on 5700 trials. During training sessions where his accuracy was above 80%, he could use an auditory speller to produce one letter per minute, and freely spell intelligible sentences on 44 out of 107 days, which allowed him to express some of his needs. However, despite these encouraging results, invasive devices cannot be proposed to all patients, because the risks associated with the implantation (infection, hemorrhage) have to be compensated by the expected benefits. The truth is, potential advantages remain strikingly difficult to estimate. Indeed, as we saw previously, when facing patients with complete paralysis, there is a huge uncertainty about their consciousness and their cognitive abilities. In this context, non-invasive BCI could help detecting to patients with residual voluntary mental activity and provide them with a first line communication tool.

When targeting patients who, by definition, cannot use motor control, including oculomotor one, non-visual BCI have to be considered. Some translational studies suggest that the auditory modality could provide a way to reach these patients [14]–[17]. In these four studies, one used pure tones as stimuli, which required the patients to learn a “code” (there were two different frequency streams, one standing for “yes” and the other one for “no”) [15]. This kind of code is quite difficult for patients with possible memory impairment. That’s why other authors suggested the use of spoken words. Sellers et al. [16] and Lulé et al. [14] proposed an oddball protocol where the four words “yes”, “no”, “stop”, “go” were delivered in a random order. The patients with LIS or with disorders of consciousness were asked to count the target words, in order to elicit a P300 event-related potential. In Lulé et al. [14], one out of two persons with a LIS could control the BCI with an online accuracy of 60%. An offline analysis showed that one patient with disorders of consciousness had 8 correct answers out of 14. The only signal considered for classification was the P300 response to deviant stimuli, thus neglecting the potential information carried by responses to standard sounds. A study by Hill et al. [18] showed that it was possible to further use the attentional modulations of the N200 wave elicited by standard words (“yes” and “no”), on top of the ones associated with deviant stimuli. They obtained a fairly good binary classification with healthy subjects (77 % +/-11 s.e. with 100 trials, chance level ∼62 % with an alpha risk of 1 %). They tested this paradigm in two ALS patients at an advanced stage of the pathology, and obtained accuracies comparable to the ones in healthy subjects.

Considering these encouraging results, we implemented and tested a “yes/no” auditory-based BCI exploiting the attentional modulations of responses to both standard and deviant sounds. We first assessed the auditory BCI with healthy controls. Then, we tested it with a group of 7 patients with severe motor disability but with residual means of communication. This enabled us to (1) be sure that instructions were perceived and understood, (2) get feedback and adapt the paradigm to each patient whenever needed to maximize our chance of success. This is important at this stage, as gaze-independent BCI have rarely been tested in patients so far.

## Methods

We used an oddball paradigm, expecting a salient P300 like response to (duration) deviant stimuli when subjects are voluntarily paying attention to them. We also expected an attentional effect onto responses to standard sounds, for the attended stream of stimuli. Regarding further analysis, we computed offline performances by considering less stimuli/block, less electrodes or different preprocessing pipelines. We also assessed the evoked potentials, both at the group and individual level, in order to identify the electrophysiological responses associated with BCI control.

### Experimental design

#### General presentation

We used spoken words pronounced by a synthetized male voice (“yes” and “no”). The sound duration was 100 ms for standard against 150 ms for deviant sounds. The stimulus onset asynchrony (SOA) was set to 250 ms for healthy subjects and adjusted to each patient (*Table 1*). The audio file of the stream of stimuli is provided in supplementary material (*Audio file 1*).

**Table 1:** Experimental conditions for each patient.

| PATIENTS | SET UP | NUMBER OF ELECTRODES | LATERALIZED STIMULATIONS<br>("YES" ON THE RIGHT,<br>"NO" ON THE LEFT) | CALIBRATION(S) |  |  |  | TEST |  |
| --- | --- | --- | --- | --- | --- | --- | --- | --- | --- |
|  |  |  |  | 1<sup>ST</sup> |  | 2<sup>ND</sup> |  | SOA<br>(ms) | Number<br>of blocks |
|  |  |  |  | SOA<br>(ms) | Number<br>of blocks | SOA<br>(ms) | Number<br>of blocks |  |  |
| LIS1 | Vamp™ | 16 | No<sup>c</sup> | 350 | 14 | 350 | 14 | NA | NA |
| LIS2 | Vamp™ | 16 | Yes | 350 | 14 | 350 | 14 | NA | NA |
| LIS3 | Vamp™ | 16 | Yes | 350 | 7<sup>a</sup> | 500 | 14 | 500 | 20 |
| ALS1 | BrainAmp™ | 32 | Yes | 300 | 14 | NA | NA | 300 | 30 |
| ALS2 | BrainAmp™ | 27<sup>b</sup> | Yes | 300 | 14 | 400 | 14 | 400 | 20 |
| ALS3 | BrainAmp™ | 32 | Yes | 400 | 14 | NA | NA | 400 | 30 |
| ALS4 | BrainAmp™ | 32 | Yes | 400 | 14 | NA | NA | 400 | 30 |

**Legend:** A calibration was usually based on 14 blocks and tests were repeated by bunchs of 10 blocks, but only if the patient was willing to pursue. The SOA was adjusted if the patient told us that the stimulations were presented at too high a rate. We usually began with a SOA of 300 ms.
NA: Not available
<sup>a</sup> Shortened calibration because the patient told us it was going too fast
<sup>b</sup> No electrode at the back of the head because of huge artifacts
<sup>c</sup> Unilateral stimulations because this patient was deaf in one ear (unilateral deafness that existed prior to the brainstem lesion).

The “yes” sounds were delivered on the right ear and the “no” sounds on the left ear (*Figure 1*). The two streams were intermixed, meaning that the “yes” and “no” sounds were never presented at the same time. The “yes” stream always started 250 ms before the “no” stream.

**Figure 1:**
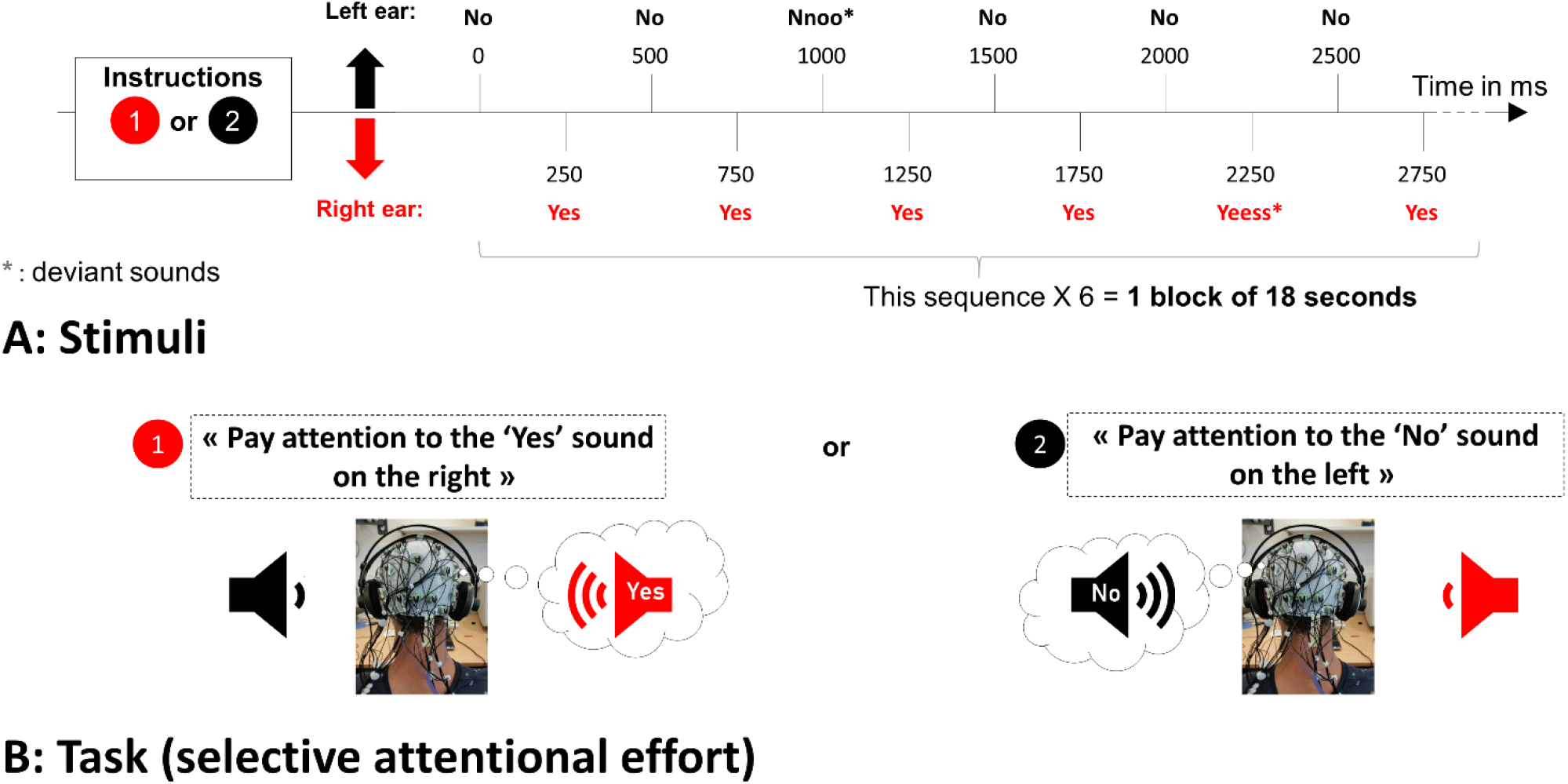
Auditory brain-computer interface protocol for healthy subjects (SOA = 250 ms). One block comprised 30 standards sounds and 6 deviants of each category (“Yes” or “No”), i.e. 72 stimuli. The deviant sounds happen randomly. It took 18 seconds to obtain an answer.

There were 30 standard and 6 deviant sounds per block, for each stream (i.e. 6 “yes” and 6 “no” deviants, respectively), yielding 18-second long block in healthy subjects, and up to 36-second long ones in patients. The deviant and standards were presented in a pseudo-randomized order, with at least two standards in-between two deviants. Users were asked to focus their attention on one stream only (the “ATTENDED” stream), and to count the number of deviant sounds in that stream. For example, to convey a “yes” answer, the subjects had to focus on the right, pay attention to all “yes” speech sounds and to count the number of “yes” deviant. At the same time, they had to ignore the concurrent stream (the “IGNORED” stream). Stimuli were delivered through headphones (*Figure 1*). The volume was standardized around 75 dB for all participants. The sounds were sent using the “NBS Presentation®” software, controlled by a script written in Python which also processed the EEG signal online.

The EEG system was an Acticap™ (Brainproduct) with 32 active gel-electrodes (*Figure 2*). The international 10–20 system was used for electrode placement, with the ground electrode on the forehead and the reference on the nose. The signal was sent to a BrainAmp™ amplifier and digitized at 1000 Hz for healthy subjects, or to a V-Amp™ amplifier (BrainVision) for the first three patients (with a sampling frequency also at 1000 Hz, and the same type of gel-electrodes). We used 16 electrodes for the first three patients, with the set up shown in Figure 2. Then considering the poor online performance in these patients, we decided to go back to a 32-channel set-up, as used with healthy subjects, for the last 4 patients.

**Figure 2:**
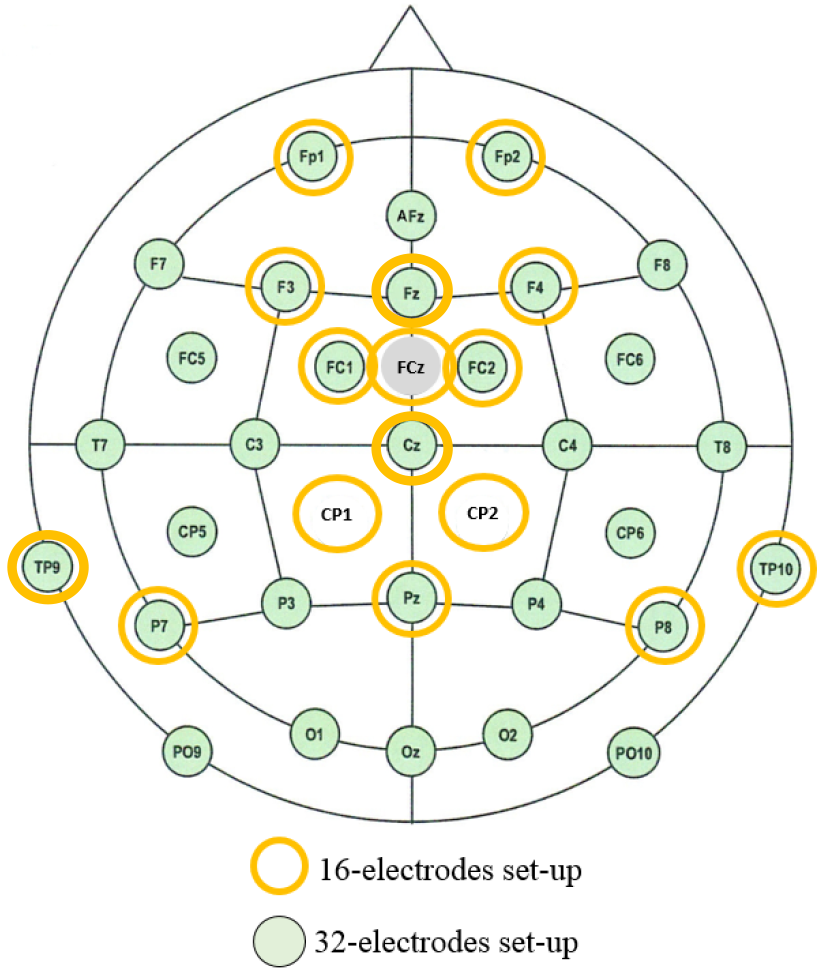
EEG electrode lay-out

#### Online signal processing

A band-pass filter between 0.5 and 20 Hz was applied. Spatial filtering, computed by the xDAWN algorithm [19], was then used to reduce dimensions and maximize the distinction between the “ATTENDED” and “IGNORED” responses. xDAWN yields as many spatial filters (or virtual sensors) as the number of EEG sensors. Importantly, spatial filters are orthogonal to each other and ranked according to how much they separate the two signals. We used between 1 and 5 filters. The exact number was optimized for each subject based on the results of a leave one out cross-validation procedure performed on the calibration data.

In healthy subjects, 500 ms and 750 ms long epochs were considered to analyze the responses evoked by standard and deviant sounds, respectively. In patients, considering that they often have delayed event related potentials (ERPs), we considered larger windows, namely 800 ms for standard and 1000 ms for deviant sounds.

Averaging was performed for each of the four conditions: standard “yes”, standard “no”, deviant “yes” and deviant “no”. A Bayesian classifier computed the posterior probability of each class given the observed features (“yes” or “no” target stimuli versus “yes” or “no” non-target stimuli, for both standard and deviant sounds, respectively). This means that one classifier was trained and used for each binary classification, using calibration data. Ultimately, the outputs of each of the four classifiers were optimally combined to obtain a final and unique posterior probability for each of the two classes (“yes” or “no”).

Right after the calibration phase (14 blocks), a leave-one-out cross-validation procedure was used to evaluate the quality of this calibration. The cross-validation is made by block (i.e. including 30 standards and 6 deviants “yes” and the same amounts of “no”). Depending on the result, it could be decided to proceed with the online testing of the BCI or to perform another calibration if performance were no better than chance.

### Healthy subjects

The study was approved by an ethical committee (Ancillary project included in trial N° NCT03233282). Within a single session, 19 healthy subjects (10 females, 23.2 ± 4.4 years) had to perform a 14 blocks long calibration, where responses to provide were instructed, followed by 50 very simple open questions for testing. The questions were printed on screen. Fourteen subjects were complete naive users while 5 subjects (n° 1, 2, 16, 17 and 18) had already taken part to one fairly similar auditory BCI experiment about a month before. Only subject n° 2 had to be excluded because it turned out that he did not perform the task as instructed to.

### Patients with severe motor impairment

Seven patients were recruited at the University Hospitals of Lyon and Saint-Etienne, in rehabilitation wards and reference centers for ALS. We informed the patients and their legal representatives about the protocol. After that, patients gave their agreement with a “yes-no” motor code, and for each of them, as they couldn’t write, their legal representative gave a written consent. The study was approved by the ethical committee Sud-Est III (Clinical trial N° NCT02567201).

Clinical information for each patient is reported in *Table* 2.

**Table 2:** Patients’ clinical characteristics

| N° | PATHOLOGY | ETIOLOGY (LIS) OR ONSET (ALS) | AGE (RANGE OF YEARS) | PATHOLOGY DURATION | COMMUNICATION CODE | ALS-FR SCALE |
| --- | --- | --- | --- | --- | --- | --- |
| LIS1<sup>A</sup> | LIS | Hemorrhagic stroke in the pons | 50-59 | 3 years | Yes-No code with vertical eyes movements, alphabetic code, no high-tech devices | NA |
| LIS2 | LIS | Traumatic with brainstem injury | 30-39 | 3 years | Yes-No code with vertical eyes movements, alphabetic code, no high-tech devices | NA |
| LIS3 | LIS | Ischemia (post-traumatic) with brainstem injury | 60-69 | 32 years | Yes-No code with vertical eyes movements, alphabetic code, eye-tracker and chin contactor | NA |
| ALS1 | ALS | Bulbar onset | 50-59 | 6 years | Yes-No code with head movements, chin contactor | 17/48 |
| ALS2 | ALS | Limbs onset | 30-39 | 5 years | Yes-No code with eyes-movements with a fluctuant reliability, eye-tracker at test | 0/48 |
| ALS3 | ALS | Limbs onset | 50-59 | 4 years | Yes-No code with head movements, alphabetic code by laser pointing with head, no high-tech devices | 18/48 |
| ALS4 | ALS | Limbs onset | 60-69 | 25 years | Phonation (interpretation with help of caregiver), eye-tracker | 23/48 |

**Legend:** ALS: Amyotrophic Lateral Sclerosis; ALS-FR Scale: ALS Functional Rating Scale; LIS: Locked-in syndrome;
A) Patient LIS1 presented a unilateral deafness (pre-existing before the brainstem injury), so he underwent a particular protocol with no lateralization of the stimulations.

The clinical investigators of the reference centers for ALS assessed the patients with the ALS Functional Rating Scale [20]. Each patient underwent a single recording session. To limit the cognitive load, we did not use questions but only direct instructions. For example, for healthy subjects, we asked “Is Paris the capital of France?”. Whereas for patients we asked “Pay attention to the right side, trying to count the “yes” that are longer” (see *Figure 1* for an example). Applying some user-centered design principles [21], we individualized the protocol for each patient, taking into account their feedback on the workload during the experiment, especially considering the speed of the streams, which led to different SOA between patients. A calibration was usually based on 14 blocks and tests were repeated by bunch of 10 blocks, but only if the patient was willing to pursue. The SOA was increased if the patient complained that the pace of stimulus presentation was too fast, starting with 300 ms.

Standardized written instructions were given aloud by the experimenters to introduce the experimental protocol. We presented sequences of stimuli to the patients prior to the actual experiment, especially to highlight the difference between standard and deviant sounds. In this initial familiarization phase, all patients were asked to count the deviant sounds and to report the number, in order to check that they were able to detect them. As already mentioned, we adjusted the experimental procedure according to the results of calibration. If performance, as estimated by the cross-validation procedure, was at chance level, a new calibration was performed. Otherwise, a test was launched during which the patient was receiving online auditory feedback on her BCI performance (e.g: “The selected answer is yes”). The test was made of bunch of ten consecutive blocks. Pauses between blocks allowed us to check the patient’s state of comfort and fatigue. We would also briefly interrupt the experiment if the patient needed respiratory care or felt uncomfortable for some other reason.

In the rare event that the performance of the second calibration was also at chance level but the patient was eager to try the BCI mode, we decided to go into test mode anyway, so as not to lengthen the experiment further with a new calibration, while hoping to obtain enough data to reliably evaluate *a posteriori*, offline, the overall performance of this patient. *Table 2* summarizes the experimental conditions experienced by each patient.

### Offline analysis (patients and controls)

#### Brain-Computer Interface offline simulation

Complementary offline analyses were performed to assess and predict BCI performance based on different numbers of spatial filters and different numbers of accumulated evoked responses per block. For sake of homogeneity, these offline analyses were based on the 15 electrodes set-up that all subjects (controls and patients) had in common, and the same number of spatial filters (n=2, which was optimal for most of the subjects). We also compared the classification accuracies based on one, several or all evoked response types combined. For example, we especially compare the accuracies between classification based on responses to the “yes” and “no”, respectively, to assess if their different acoustical properties had an impact. This was done with the same classifier used online. We also assessed whether the classification result would be different if independent component analysis (ICA) would be used to remove artifacts (blinks, saccades and artefacts on reference electrode).

At the end of the control study, before the clinical trial, an offline procedure was performed in order to select the most relevant sensors. We used a backward selection. We first computed the accuracies obtained with all electrodes, and then removed step-by-step the least contributing one in order to identify the most informative sensors at the group level. This analysis was motived by the practical aim of possibly optimizing our set-up, making it more portable and faster to install for our clinical tests in different clinical centers.

#### Processing of the evoked potentials

The MNE python software [22] enabled us to analyze evoked related potentials (ERP). Raw EEG data were pre-processed by ICA to remove artifacts due to eye movements (blinks and saccades) and common artifacts due to disturbance on the reference electrodes. The pre-processed data were then filtered with the same band-pass filter as used online (0.5-20 Hz). For each stimulus type (standard “yes” and “no” and deviant “yes” and “no”) and condition (target and non-target), single responses were epoched between –200 ms to +1000 ms with respect to stimulus onset. There was a rejection of 15% of the epochs based on their amplitude. No baseline correction was applied, both because of the use of a short SOA and to conform to the processing steps used online. For each subject, we merged the calibration and test data. In controls, we also computed the grand average ERP for each condition, only including subjects who performed best in controlling the BCI (n= 11, online accuracy > 88%), to better characterize the biomarkers of a good BCI control. We also performed an individual ERP analysis for each subject. We looked at the presence of an attentional modulation, namely a significant difference between the “ATTENDED” and the “IGNORED” sounds. We further analyzed the presence of a specific evoked response to deviance compared to standards.

We hypothesized that some subjects could use, even involuntarily, their eyes or their eyelids to control the BCI. As we did not record directly the electro-oculogram, we used indirect markers to assess this hypothesis. We extracted the ICA components of saccades and blinks. For each component, we averaged separately the “ATTENDED” and “IGNORED” stimuli, and we performed a statistical test to check if there was an attentional modulation of these ICA components. In other words, the ICA was used here as a spatial filter on the sensors that were the most sensitive to the ocular and eyelids artefacts.

### Statistical analysis

#### BCI performance: Accuracy and chance level threshold

Our primary criterion was BCI performance, assessed by classification accuracy, that is, the percentage of blocks for which the selected response was correctly identified. An accuracy below chance level was interpreted as a complete lack of BCI control. In order to test for the statistical significance of the obtained accuracies, we assumed that classification errors obey a binomial cumulative distribution, as described in [23]. Therefore, the empirical chance level depends on the total number of blocks. We performed this comparison with accuracies obtained with the 15-channels and 2 filters used for the offline analysis.

#### Statistical comparison of evoked potentials

Both at the group (grand average) and at the individual level, we compared the attended versus ignored stimuli. We used a non-parametric cluster-level test for spatio-temporal data [24], with a threshold-free cluster enhancement [25], 10,000 permutations and a p-value threshold of 0.01. For the comparison of attended versus ignored evoked responses of the ICA components, we performed a cluster-level statistical permutation test with 10000 permutations and a p-value threshold of 0.05.

## Results

### Healthy subjects

#### Online results

The online accuracy obtained at the group level was 86 % on average (n= 18, s.e: 11.7%, median: 92%). Ten subjects obtained an accuracy above 90 %. For the 14 naive subjects, the mean accuracy was also 86%.

#### Offline results

The offline performance was similar with and without ICA preprocessing (*Figure 3*).

**Figure 3:**
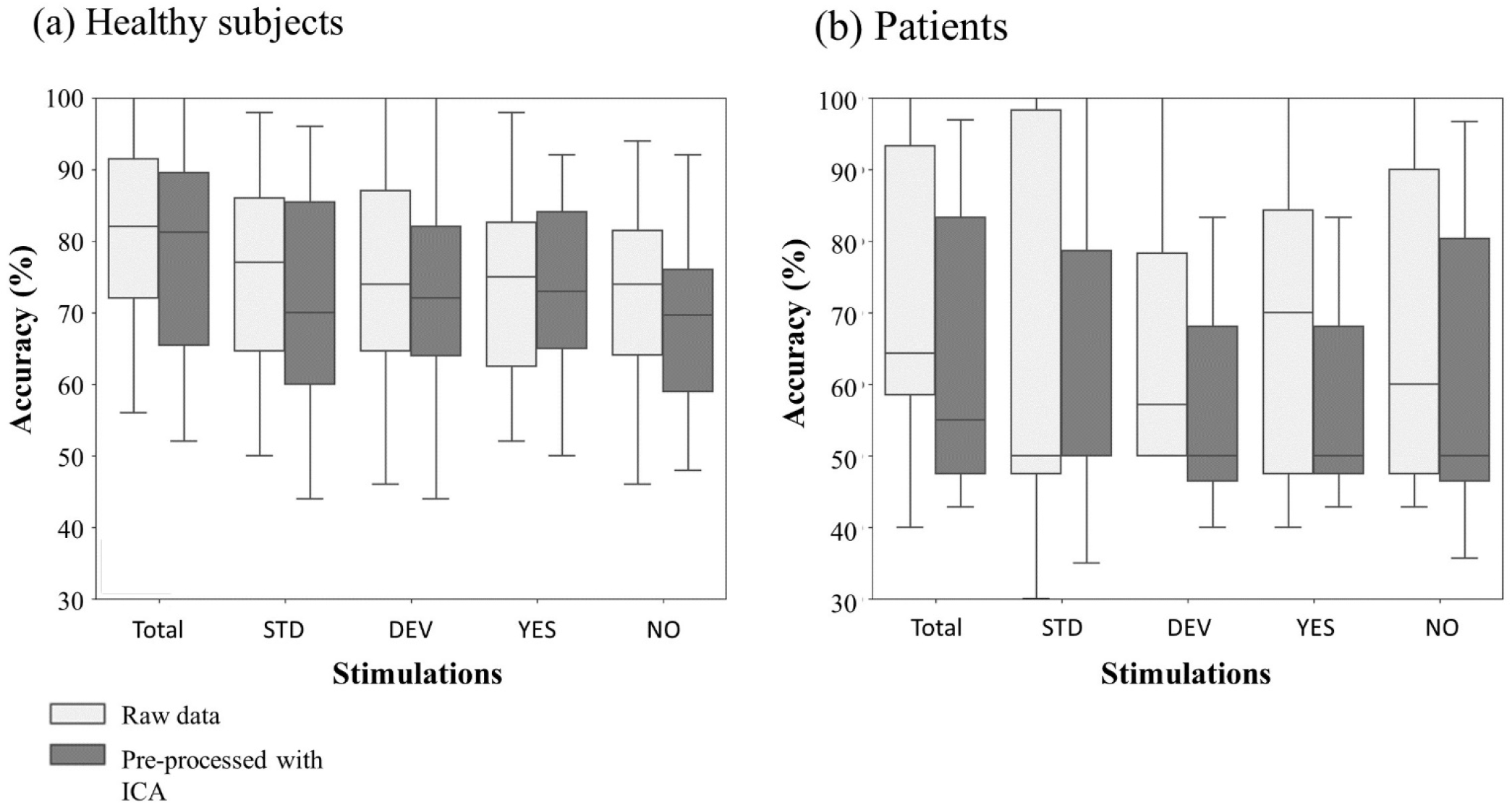
Offline classification results with and without ICA correction. Offline BCI simulations with 15 channels, for healthy subjects **(a)** and patients **(b)**. Boxplots filled with light gray stand for accuracy results with no pre-processing of the data, except filtering. Boxplots filled with dark gray stand for the condition where the signal was preprocessed with ICA, removing blinks, saccades and a DC component. **Stimulations: Total:** Pool of responses to all stimuli, **DEV:** Pool of all responses to deviants, **YES:** Pool of all responses to « yes », **NO:** Pool of all responses to « no ».

We reanalyzed the group data (n=18, 50 blocks each) for various block lengths. We observed that the accuracy remained stable for block duration comprised between 9 and 18 seconds, which is promising for future improvements of the paradigm, as it would allow to improve the information transfer rate (ITR) and would require a shorter attentional effort (*Figure 4*). We also assessed the classification performance when considering not all stimulus types together, but either standard or deviant sounds, respectively (*Figure 4*).

**Figure 4:**
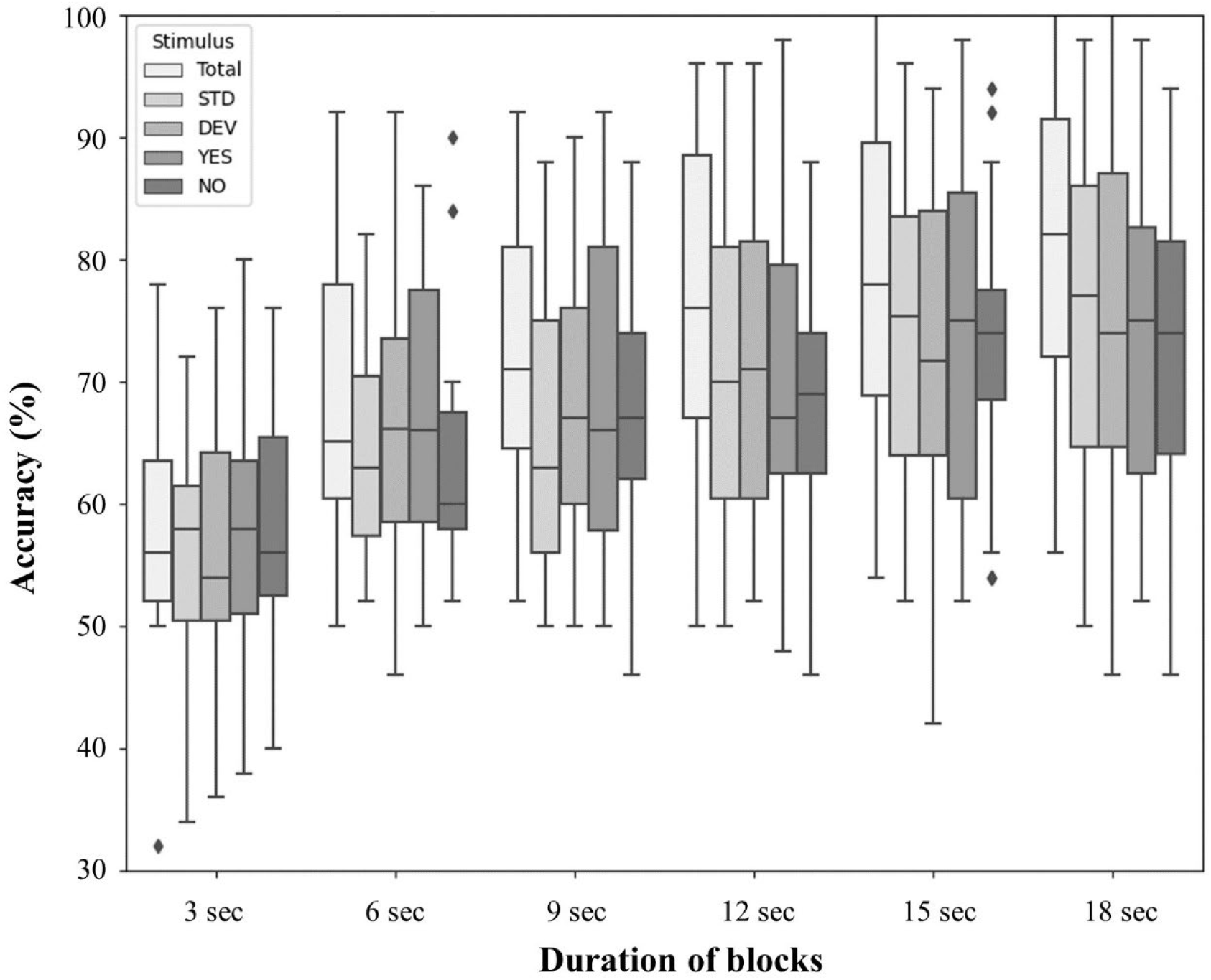
Offline classification results with different duration of blocks. Offline BCI simulations with 15 channels, for healthy subjects. **Stimulations: Total:** Pool of responses to all stimuli, **DEV:** Pool of all responses to deviants, **YES:** Pool of all responses to « yes », **NO:** Pool of all responses to « no ».

It appears that the best accuracies were obtained when accounting for the combination of standard and deviant responses, compared with accuracies based on standards or deviants only. We did not find a significant difference between yes and no responses, despite their different acoustic properties. Finally, the backward electrode selection procedure revealed that performance remained unaffected when reducing the EEG set down to 15 sensors (*Figure 5*).

**Figure 5:**
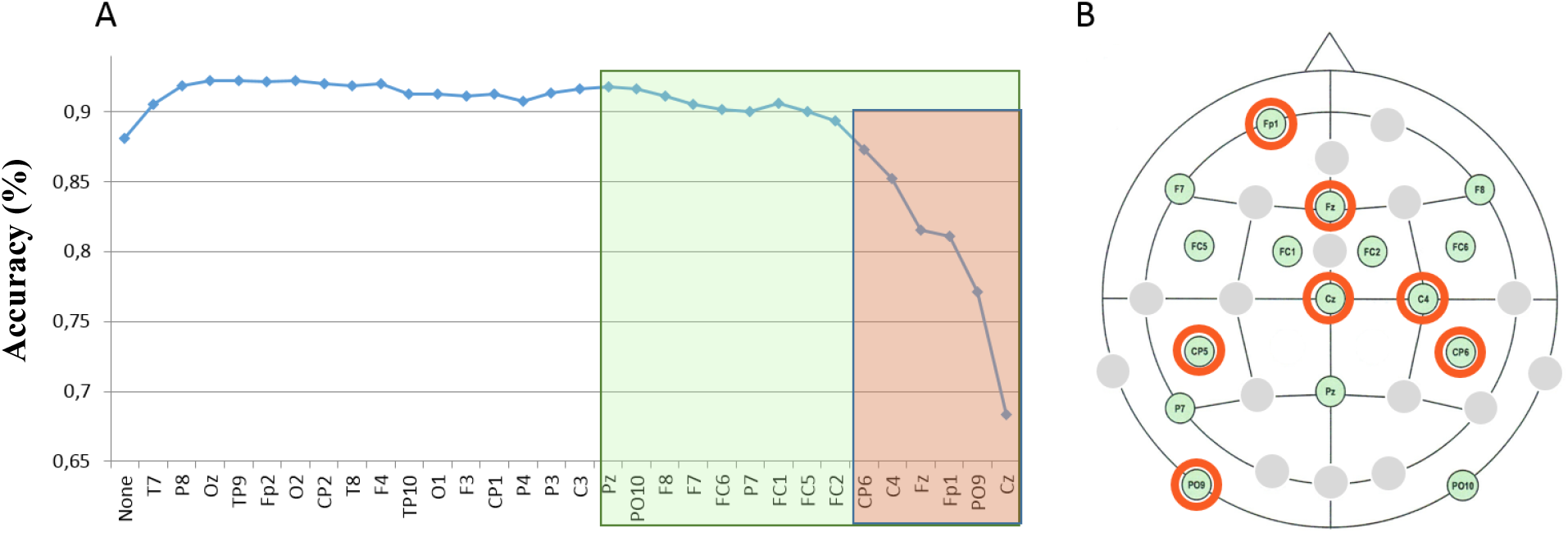
Backward selection of relevant electrodes. (A) Horizontal axis, from left to right: at each step of the backward selection, an additional electrode is removed (the one that minimizes the loss in accuracy). Vertical axis: Accuracy. (B) Spatial locations of the most relevant 16 (resp. 7) electrodes (green resp. red rectangle).

This suggest that future experiments with healthy subjects could be optimized in using a more portable device (e.g. Vamp® system) with 15 channels only (as we did for the first patients, see *Figure 2*).

#### Evoked potentials

Average responses to standards, for the group of subjects who best controlled the BCI (n=11, accuracy > 88 %), revealed a positive peak at 65 ms and a negative one at 115 ms after stimulus onset. The latter is reminiscent of the auditory N1 component (*Figure 6a*). At the group level, there was no attentional modulation on standard stimuli. However, at the individual level, 83 % of the subjects showed an attentional modulation on standards. This modulation was highly variable in terms of latency and duration. For the deviant response, we observed a clear P3a component (*Figures 6b*), followed by a large P3b when the subject paid attention to the target.

**Figure 6:**
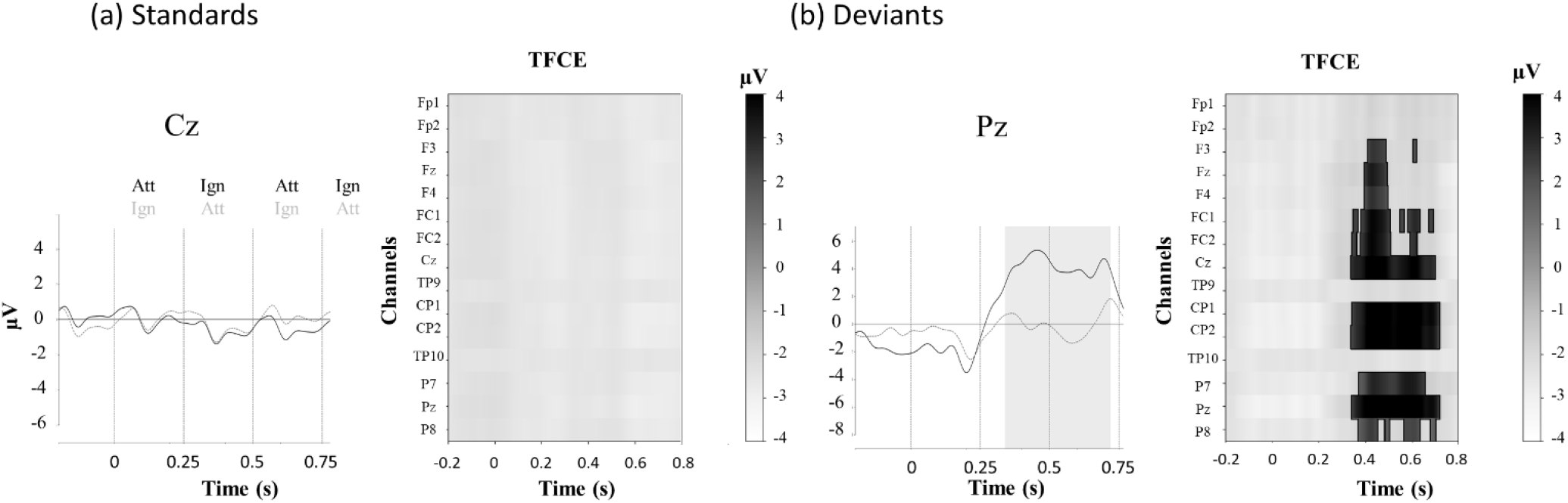
Effect of attention on event related potentials (ERPs): group average for healthy subjects. **Mean ERPs** for attended and unattended sounds, standards (a) and deviants (b). The SOA is at 250 ms, with an alternance of ATTENDED (Att) and IGNORED (Ign) sounds, hence a switch in the attentional modulation every 250 ms. Each stimulus onset is represented by a vertical dashed line. The shaded area corresponds to the period when this difference is significant. This analysis was performed on the preprocessed signals using ICA. **TFCE:** Threshold-free cluster enhancement test for the difference between attended and ignored sounds. Each line represents one electrode. When significant, the clusters for one electrode appear in white (negative) or in gray (positive). There is no significant cluster for the standards.

At the individual level, 72% of healthy subjects had an attentional modulation on deviants. Ninety-four percent of the subjects had a response to deviance (*Table 4*).

### Patients with severe motor impairment

#### Online results

All patients could hear at least some of the deviant sounds. However, 5 out of 7 patients could not control the BCI: online performance was at chance level or below (*Table 3*). The other two patients (ALS3 and ALS4) did achieve a high degree of control of the BCI, with a 100% accuracy over 30 blocks.

**Table 3:**
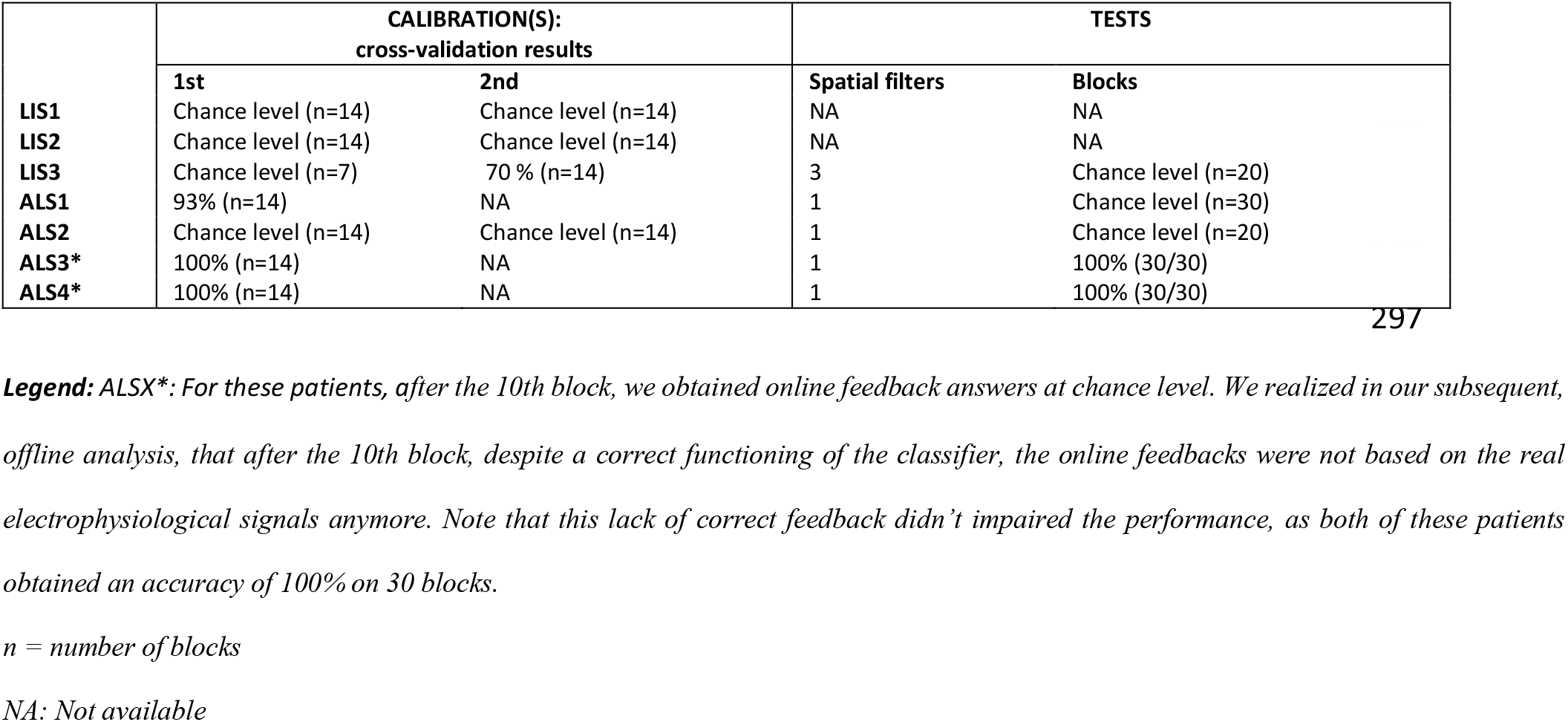
Patients’ online results

#### Offline results

With a 15-channel set-up, patient ALS1 who performed at chance level online, improved offline, up to 87 % accuracy. Patients ALS3 and ALS4, who performed with 100 % accuracy online, with 32 electrodes, maintained their performance when considering those 15 electrodes only (97 % and 100 % respectively). Interestingly, the offline BCI simulation with ICA preprocessing (*Figure 3b*) did not yield any difference compared to raw data, except for patient ALS1 whose performance in session 2 dropped by 24 % (from 87 % to 63 %). We observed that relying on part of the data (e.g. deviants or standards only) yields similar performance than when considering all the data.

#### Evoked potentials

Individual analysis revealed an artefacted signal in all patients compared to healthy subjects. There were different kinds of artefacts. In one patient (LIS2), it was clearly due to the pathology: he had a facial spasm that contaminated all channels and prevented the visualization of evoked potentials, which did not allow a good functioning of the BCI either. In other patients, it was more electrical artifacts likely due to mechanical ventilation or to the pump of the inflatable air mattress used in prevention of bed sores. However, in the latter cases, signal preprocessing allowed the visualization of evoked potentials.

A significant attentional modulation of the evoked potentials could still be observed in patients who managed to control the BCI (ALS3 and 4) for both standards and deviants (*Figure 7*).

**Figure 7:**
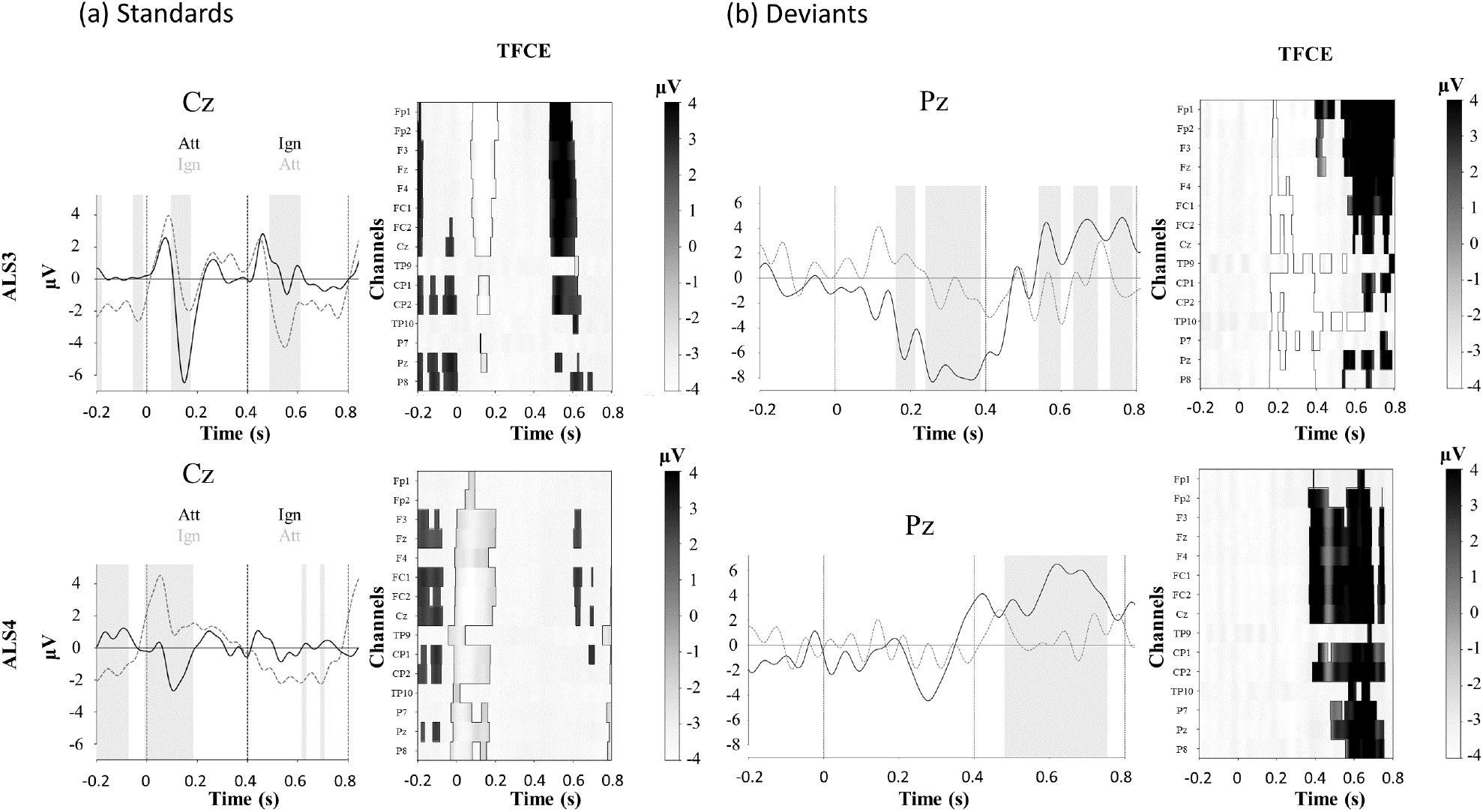
Effect of attention on averaged evoked related potentials (ERPs) to sounds “yes” in patient ALS3 and ALS4. Mean ERPs for attended and unattended sounds, standards (a) and deviants (b). The SOA is at 250 ms, with an alternance of ATTENDED (Att) and IGNORED (Ign) sounds, hence a switch in the attentional modulation every 250 ms. Each stimulus onset is represented by a vertical dashed line. The shaded area corresponds to the period when this difference is significant. This analysis was performed on the preprocessed signals using ICA. **TFCE:** Threshold-free cluster enhancement test for the difference between attended and ignored sounds. Each line represents one electrode. When significant, the clusters for one electrode appear in white (negative) or in gray (positive).

As for healthy subjects, the pattern of attentional modulations for standards varied from one patient to the next in terms of latency and topography. *Figure 7* shows an example of this attentional modulation in response to “yes” standards in patients ALS3 and ALS4. The topography of the significant difference between the attended and unattended conditions is mainly fronto-central. It was maximal at frontal level without ICA pre-processing. After removing the ICA components corresponding to blinks, saccades and a common offset, it was still present in all electrodes. Amongst patients showing no BCI control, they were no attentional modulation at the classical localization of the evoked responses (central and parietal electrodes).

Albeit all patients could behaviorally detect at least some deviant sounds, and contrary to what we observed in most of the healthy subjects, five out of seven patients did not show a significant response to deviance (*Table 4*). Patient ALS4 presented with a P300 response to attended deviants, while patient ALS3 also showed a response to attended deviants around 300 ms, but with a negative polarity (*Figure 7*).

**Table 4:** Controls and patients’ event-related potentials

| Subjects | Online accuracy | Response to deviance | Attentional modulation |  |  |
| --- | --- | --- | --- | --- | --- |
|  |  |  | Standards | Deviants | Total |
| S1 | 100 | ✓ | ✓ | ✓ | ✓ |
| S3 | 68 | ✓ | Ø | Ø | Ø |
| S4 | 80 | ✓ | ✓ | ✓ | ✓ |
| S5 | 88 | ✓ | ✓ | Ø | ✓ |
| S6 | 92 | ✓ | ✓ | ✓ | ✓ |
| S7 | 92 | ✓ | ✓ | ✓ | ✓ |
| S8 | 96 | ✓ | ✓ | ✓ | ✓ |
| S9 | 82 | ✓ | ✓ | ✓ | ✓ |
| S10 | 96 | ✓ | ✓ | ✓ | ✓ |
| S11 | 96 | ✓ | ✓ | Ø | ✓ |
| S12 | 72 | ✓ | ✓ | ✓ | ✓ |
| S13 | 94 | ✓ | ✓ | ✓ | ✓ |
| S14 | 80 | ✓ | ✓ | Ø | ✓ |
| S15 | 60 | ✓ | Ø | ✓ | ✓ |
| S16 | 96 | ✓ | ✓ | ✓ | ✓ |
| S17 | 94 | ✓ | ✓ | ✓ | ✓ |
| S18 | 96 | ✓ | ✓ | ✓ | ✓ |
| S19 | 74 | Ø | Ø | Ø | Ø |
| **Prevalence of healthy subjects with BCI control** | **94 %** | **94 %** | **83 %** | **72 %** | **89 %** |
| LIS1 | Chance level | Ø | Ø | Ø | Ø |
| LIS2 | Chance level | Ø | Ø | Ø | Ø |
| LIS3 | Chance level | Ø | Ø | Ø | Ø |
| ALS1 | Chance level | Ø | Ø | Ø | Ø |
| ALS2 | Chance level | Ø | Ø | Ø | Ø |
| ALS3 | 100 | ✓ | ✓ | ✓ | ✓ |
| ALS4 | 100 | ✓ | ✓ | ✓ | ✓ |
| **Prevalence of patients with BCI control** | **29%** | **29%** | **29%** | **29%** | **29%** |

**Legend:** ✓: presence Ø: absence
**Response to deviance:** presence or absence of a significant difference of event-related potential for standards versus deviant sounds.
**Attentional modulation:** presence or absence of significant difference of event-related potential for attended versus ignored condition.

The analysis of each ICA component did not reveal that the attentional modulation was more captured by one or a few components compared to others. Interestingly though, some differences appeared in the ICA components between the group that controlled the BCI (healthy subjects and patients) and the patients that did not control the BCI. In the group with a good control, all subjects presented with a typical saccadic component, indicating the usual presence of oculomotor movements. However, in the group that couldn’t control the BCI, we found no obvious saccadic ICA component in three out of the 5 patients. The latter suggests that those patients had a more severe oculomotor impairment.

## Discussion

Seventeen of 18 healthy subjects proved able to control the proposed auditory BCI. In contrast, only 2 out of 7 severely motor impaired patients proved able to control the interface online, and 3 out of 7 after careful offline signal processing.

The analysis of deviant evoked responses revealed that the presence of a classical P300 and its attentional modulation was associated with a good control of the interface, in both healthy subjects and patients. This could explain the poor BCI results observed in most patients, for whom no P300 was detected. Other studies uncovered this lower prevalence of P300 in patients with LIS [26]. However, this lack of P300 response is quite surprising, since all patients could hear at least some of the deviant sounds when presented with the different stimuli. Hence this oddball auditory protocol lacks robustness with patients with severe motor impairment, and relying only on deviant sounds would not allow an accurate enough BCI communication.

For standard stimuli, the effect of attention on evoked potentials is reminiscent of an “attentional phase shift”, similar to the one observed in [27], [28]. This attentional phase shift was robust and present in 15 of the 18 healthy subjects. However, it was not visible at the group level because of a phase difference in the shift from one subject to another, a variability which is also described in [28]. This attentional shift or marker of sustained attention orienting was also present in patients who did control the BCI (*Figure 7*).

We observed no obvious N100 evoked potential at the group level. This could be explained by the variability of the evoked potentials when using words as stimuli instead of sharp tones, as noticed by Hill et al [29]. Peak latencies then indeed vary a lot between, as well as within subjects.

An important finding is that patients with severe motor disability, although clearly conscious and with residual means of communication, present with poor performance of BCI control. Only 3 out of 7 patients were able to control the BCI with an accuracy above chance level. Together with our offline analysis of their electrophysiological responses, this suggest that BCIs that are validated in healthy subjects are unfortunately not readily usable by the targeted end users. Baring in mind that, in the long term, such interfaces are mostly meant to help people who have no means of communication, our findings raise crucial challenges for our community. Reasons behind the poor BCI performance in the majority of the patients have to be thoroughly explored in order to come up with efficient non-invasive solutions.

We can put forward several, non-mutually exclusive, hypotheses. First, the quality of the signal is, on average, lower in patients (due to several factors: mechanical ventilation, erratic muscle activity, electrical interference due to hospital beds, etc.). Second, there is more and more evidence that motor impairments come with cognitive impairments, whatever the etiology [30]. In this context, it might be that our paradigm is cognitively too demanding for the patients: binaural listening requires not only focusing on the “ATTENDED” stream, but also inhibiting the “IGNORED” one. In addition to that, patients have to be able to understand fairly complex instructions, and sustain their attention for half an hour or so. However, all the patients included in this study could handle the complexity of communicating with a yes-no code using a letter board, which presupposes the preservation of some cognitive abilities, especially in terms of working memory and executive functions. Despite this ability, more than half of them proved enabled to control the BCI.

It seems difficult to further simplify the protocol given the intrinsic and technical limitations of EEG [30]. However, one possibly useful change to be tested would be to no longer present the “yes” and “no” streams concurrently, but alternately in the form of short separate blocks. This non-lateralized paradigm could be useful also for patients with unilateral deafness. This approach could make the attentional task easier, without too much extending the duration of a block. Patients would concentrate on sounds during blocks where the relevant answer is presented, while during irrelevant blocks they would divert their attention away from sounds (e.g. by imagining navigating in a familiar environment [31]). This may reduce the mental workload and could help patients with cognitive impairments, especially frontal ones, which are quite frequent at an advanced stage of ALS and can occur in LIS too. Moreover, some studies suggest that it is possible for persons with motor impairment to improve their BCI performance with training over several sessions [32].

Beyond improving the protocols, there is a need for better understanding the particularities of patients with severe motor impairment, which remain poorly explored at the moment, both at the neurophysiological and cognitive level [33]. Here we chose an auditory protocol to overcome the oculomotor limitations of patients with severe motor impairment. Indeed, oculomotor impairments are known to be a predictor of weak control of visual BCIs [34], even when stimuli are all presented at the same place in an SSVEP paradigm [35]. A recent study with audio-visual stimulations also reported chance level accuracy with a patient in CLIS (no voluntary control of eye movements), despite the possibility to detect, offline, some differences between target and non-target responses, suggesting that the patient did try to do the task [36].

In the same vein, it is striking to notice that, in our study, none of the patients with “classical” LIS, who present more often with oculomotor impairments than patients with ALS [37], managed to control the BCI. And none of the patients with no ICA component reflecting saccadic activity could control the BCI, either. Concomitantly, there is a bunch of evidence in the literature that eye-movement planning and spatial attention are tightly related [38]–[40], although not completely similar [41], [42]. Most of these studies relate to visual spatial attention, but attention is a cross-modal effort: for example, orienting attention toward a tactile target also triggers an automatic displacement of spatial attention in the visual modality [43]. Hence it would be useful to test the impact of eye movements on spatial auditory attention. Future studies should provide finer clinical information regarding these patients, namely about their oculomotor limitations, their ability to turn their head and their cognitive profiles. This would help identifying those who could actually benefit from BCI, as well as identifying factors that prevent their use.

## Data Availability

All data produced in the present study are available upon reasonable request to the authors.

## Acknowledgements

We thank the people with LIS and ALS and their relatives for their active participation to this study and for their very valuable feedbacks. We thank Lesly Fornoni for her help in writing the ethical committee files. We thank Pr Jean-Christophe Antoine, Dr Emilien Bernard, and Pr Jean-Philippe Camdessanché from the university hospitals of Saint-Etienne and Lyon for their valuable feedbacks and their help in recruiting the persons with ALS. We thank Pauline Mandon for her help in experimenting with patients.

PS, JM, EM, PG and JL were funded by two grants from the Fondation pour la Recherche Médicale (FRM, DEA20140629858 & FDM201906008524); PS and JM were also funded by the French government (ANR-17-CE40-0005, MindMadeClear). AO, JM, EM and DM were funded by the Fondation pour la Recherche Médicale (FRM, ING20121226307).

The COPHY team is supported by the Labex Cortex (ANR-11-LABX-0042).

Authors do not have any competing interest to declare.

## Authors contributions statement

EM, DM and JM conceived and designed the brain-computer interface and the experimental paradigm. EM and AO implemented the brain-computer interface. PG and JL recruited the patients. PS, MF and EM performed the experiments. PS, EM, DM and JM performed the offline data analysis. PS, EM and JM wrote the manuscript. All authors reviewed the manuscript.

